# SARS-CoV-2 specific memory B-cells from individuals with diverse disease severities recognize SARS-CoV-2 variants of concern

**DOI:** 10.1101/2021.05.28.21258025

**Authors:** Zoe L. Lyski, Amanda E. Brunton, Matt I. Strnad, Peter E. Sullivan, Sarah A.R. Siegel, Fikadu G. Tafesse, Mark K. Slifka, William B. Messer

**Affiliations:** Department of Molecular Microbiology & Immunology, Oregon Health & Science University (OHSU), Portland, OR 97239, USA; OHSU-PSU School of Public Health, Portland, OR 97239, USA; Division of Neuroscience, Oregon National Primate Research Center, Oregon Health & Science University, Beaverton, Oregon, USA; Department of Medicine, Division of Infectious Diseases, Oregon Health & Science University (OHSU), Portland, OR 97239, USA

**Keywords:** COVID-19, Memory B-cell, Variant of concern, limiting dilution assay, Antibody, Disease severity

## Abstract

In this investigation we examined the magnitude, breadth, and durability of SARS-CoV-2 specific antibodies in two distinct B-cell compartments: long-lived plasma cell-derived antibodies in the plasma, and peripheral memory B-cells along with their associated antibody profiles elicited after *in vitro* stimulation. We found that magnitude varied amongst individuals, but was the highest in hospitalized subjects. Variants of concern (VoC) -RBD-reactive antibodies were found in the plasma of 72% of samples in this investigation, and VoC-RBD-reactive memory B-cells were found in all but 1 subject at a single time-point. This finding, that VoC-RBD-reactive MBCs are present in the peripheral blood of all subjects including those that experienced asymptomatic or mild disease, provides a reason for optimism regarding the capacity of vaccination, prior infection, and/or both, to limit disease severity and transmission of variants of concern as they continue to arise and circulate.

## Background

Severe acute respiratory syndrome coronavirus-2 (SARS-CoV-2), the causative agent of COVID-19, emerged in 2019, resulting in 167 million cases and 3.48 million deaths worldwide (As of May 24^th^, 2021). SARS-CoV-2 infection leads to illness ranging from asymptomatic to severe, requiring hospitalization, mechanical ventilation, often leading to death [1]. SARS-CoV-2 specific antibodies (Abs) are a likely correlate of immunity and are thought to protect against repeat infection [2], and antibody mediated protection has been observed in humans, NHP studies, and in passive transfer of neutralizing Abs [2-7]. However, SARS-CoV-2 specific serum Abs decline in the months following natural infection or vaccination [3-7], and neutralizing Ab titers have been reported to be low in many convalescent, naturally infected individuals [4-8], with even lower titers against emerging SARS-CoV-2 variants of concern (VoC), calling into question both the durability and breadth of Ab-mediated protection against SARS-CoV-2.

Pathogen specific Abs secreted by plasma cells that differentiate in germinal centers in peripheral lymph nodes during and following acute infection are present in the plasma and serum. Short-lived plasma cells/plasmablasts transiently secrete Abs before undergoing apoptosis, while long-lived plasma cells (LLPCs) traffic to bone marrow and secrete Abs for months to years post-infection [5], protecting against repeat infections with homologous or closely related pathogens. Memory B-cells (MBCs) also differentiate in the germinal center and then circulate in low numbers in peripheral blood. MBCs do not secrete Abs, but instead survey the periphery for invading pathogens, poised to quickly respond to repeat infection by proliferating and differentiating into Ab secreting plasma cells/plasmablasts. MBCs have been found to respond to antigenically diverse, but related pathogens that evade preexisting, plasma Abs [6, 7]. Consequently, MBCs have the potential to play a critical role in developing host and herd immunity to SARS-CoV-2, especially in the face of waning Ab titers and the emergence of antigenic variants that differ from early lineage SARS-CoV-2.

Unease has arisen over the past several months with the emergence of specific CDC-defined [18] SARS-CoV-2 variants of concern (VoC) with mutations in the virus spike (S) protein. These variants include B.1.1.7, originating in the United Kingdom (UK), B.1.351, originating in South Africa (SA), and P.1, originating in Japan/Brazil. These mutations, particularly those in the S receptor binding domain (RBD), have been shown to increase transmissibility [13-14] as well as reduce Ab-mediated neutralization by human convalescent immune, COVID-19 vaccine sera [9, 15-19] and therapeutic monoclonal antibodies [28]. While SARS-CoV-2 specific MBCs have recently been characterized [5,8,20-22], studies assessing functional binding of MBC-derived Abs against VoC remain to be done. Such studies would answer the central question of whether the pre-existing SARS-CoV-2 specific MBC antibody repertoire can recognize and quickly respond to VoC reinfection with an Ab milieu that can either protect against or mitigate severity of VoC infection.

To address this question, we non-specifically stimulated PBMCs from a cohort of convalescent COVID-19 subjects in which the MBC fraction became antibody-secreting cells (ASCs) [8, 9] and assessed the MBC-derived Abs binding to SARS-CoV-2 parental strain WA-1 RBD and a VoC RBD containing the canonical K417N, E484K, and Y501N mutations. This longitudinal study investigates the magnitude, durability and breadth of Ab-mediated immune memory in COVID-19 immune subjects with varying disease severity ranging from asymptomatic to severe disease and provides a framework to define and predict long-lived immunity to SARS-CoV-2 and emerging variants after natural infection and/or vaccination.

## Materials and methods

### Human Research Ethics

The study was reviewed and approved by the Oregon Health & Science University Institutional Review Board (IRB# 21230). Informed consent was obtained from subjects on initiation of their participation in the study.

### Human subjects

Samples (n=24) were collected from subjects with confirmed COVID-19 infection (OHSU IRB# 21230). Forty mL of whole blood was collected for PBMCs and plasma (BD Vacutainer® Lavender Top EDTA Tubes). Whole blood was spun @ 1,000 x g and the plasma fraction was collected and stored at -80 until use. PBMCs were isolated using Sepmate tubes (Stemcell) and stored in liquid nitrogen until needed.

### Human MBC limiting dilution analysis

PBMCs were thawed and resuspended in LDA media, RPMI 1640 (Gibco), 1×Anti-Anti (Corning), 1X non-essential amino acids (HyClone), 20 mM HEPES (Thermo Scientific), 50μMβ-ME, and 10% heat-inactivated FBS (VWR). Cells were cultured in serial 2-fold diluted doses (10 wells per dose), starting with 3-5 x 10^5^ PBMCs per well at the highest dose. In 96-well round-bottom plates in a final volume of 200μl per well. Cells were incubated with IL-2 (Prospec) 1000U/ml and R848 (InvivoGen) 2.5ug/mL.[8] To determine background absorbance values, supernatants were used from 8 wells unstimulated PBMCs only. Plates were incubated at 37 °C and 5% CO^2^ for 7 days. Stimulation was determined by running total IgG ELISAs. Antigen-specific MBC frequencies were calculated by assaying LDA supernatants by antigen-specific ELISAs [9].

### Antigen-specific ELISA for plasma and LDA

Ninety-six well (plasma, Corning-3590) or half-well ELISA plates (LDA, Greiner Bio-one) were coated with 100µL (plasma) or 50µL (LDA) of 1ug/mL antigen in PBS, recombinant RBD (provided by Dr. David Johnson), recombinant spike subunit 1 (40591-V08H, Sino Biological Inc.), and recombinant VoC-RBD. Plates were incubated overnight at 4°C, washed with PBS-T (0.05% Tween) and blocked for 1 hour with 5% milk prepared in PBS-T. For plasma, samples were serially 3-fold diluted in dilution buffer and 100uL was added to wells. Plates were incubated at room temperature (RT) for 1 hour. For LDA, 20μL of LDA supernatants were added to each well and incubated at RT for 1 hour. Plates were washed 4 times with wash buffer, and 100µL (plasma) or 50 μL (LDA) of 1:3000 dilution of anti-human IgG-HRP (BD Pharmingen, 555788) detection antibody was added and incubated at RT for 1 hour. Plates were washed 4 times with wash buffer, 100µL (plasma) or 50 μL (LDA) of colorimetric detection reagent containing 0.4□mg/ml o-phenylenediamine and 0.01% hydrogen peroxide in 0.05□M citrate buffer (pH 5) were added and the reaction was stopped after 20□minutes by the addition of 1□M HCl. Optical density (OD) at 492□nm was measured using a CLARIOstar ELISA plate reader. Plasma Ab endpoint titers were set as the lowest dilution with an OD 4-fold above PBS only wells. LDA wells were scored positive at ODs at least 2-fold above background (unstimulated PBMC wells).

### MBC frequency

MBC precursor frequencies were calculated by the semi-logarithmic plot of the percent of negative cultures versus the cell dose per culture, as previously described [9] and frequencies were calculated as the reciprocal of the cell dilution at which 37% of the cultures were negative for antigen-specific IgG production. Rows which yielded 0% negative wells were excluded, since this typically resides outside of the linear range of the curve and artificially reduced the MBC precursor frequency. For subjects with low frequency of antigen-specific antibody secreting cells frequency was determined by number of positive wells divided by the total number of IgG positive secreting wells, multiplied by one million, giving a frequency per million PBMCs stimulated.

### Generation of VoC-RBD and protein expression

To generate the triple mutant VoC-RBD we started with the WT RBD gene and performed site directed mutagenesis to make the following changes: N501Y, E484K, K417N using Q5 site-directed mutagenesis kit (NEB). Purified SARS-CoV-2 VoC-RBD protein was prepared as described previously [40-41]. Briefly, His-tagged VoC-RBD bearing lentivirus was produced in HEK 293-T cells and used to infect HEK 293-F suspension cells. The suspension cells were allowed to grow for 3 days, shaking at 37°C, 8% CO2. Cells were spun down, supernatant collected, sterile filtered, and purified by Ni-NTA chromatography. The purified protein was then buffer exchanged into PBS and concentrated.

### Statistical methods

For values below the limit of detection a value of 49 was assigned for ELISA titers, and a value of <0.0000001 was assigned for MBC frequencies (corresponding to <1 SARS-CoV-2 antibody secreting cell per total PBMCs stimulated). Subjects who received vaccination prior to second draw were included in the longitudinal data, but excluded from all statistical analysis. Paired t-test was done to compare ELISA titers between RBD-WA-1 and RBD-VoC as well as used to compare log transformed specific MBC frequencies against WA-1 and VoC RBD. Spearman’s rank-order correlation was performed for all correlation analysis. All analysis was done in Prism version 9.1.1.

### Univariate and multivariate analyses

We conducted univariate analyses with each pre-determined clinical covariate of interest and MBC frequency was log transformed. Each univariate analysis was conducted as follows: age (continuous), age (categorical, <50 vs. ≥50), disease severity (ordinal), disease severity (categorical, <5 vs. ≥5), and hospitalization status (yes vs. no). For multivariable models our first model included age (continuous), disease severity score (ordinal), and the interaction between age and disease severity. Contrast estimates were evaluated for each disease severity score interaction with age to determine if any effects within each score and mean age existed. Our second model included age, hospitalization status, and the interaction between hospitalization status and age. All statistical analyses were performed in SAS version 9.4.

## Results

### COVID-19 cohort population

SARS-CoV-2 PCR positive subjects were recruited in Oregon under an OHSU IRB approved protocol (IRB# 21230). On enrollment, a comprehensive medical history was taken in addition to serum, plasma, and peripheral blood mononuclear cells (PBMC). When possible, one or more follow-up serial sample visits were arranged. Samples from 24 subjects were taken during the convalescent period (>28 days post-infection) (Table 1). Time post-infection, defined by months post-PCR diagnosis, ranged from 1-11.75 months. WHO Disease severity scores [42] (ranged from 1 (asymptomatic) to 6 (severe, requiring intubation and mechanical ventilation), with 2 asymptomatic subjects, 16 symptomatic/not hospitalized subjects, and 6 hospitalized subjects. Subjects were 46% female, with a median age of 60 (Table 1). Of the 24 subjects, two were vaccinated before their final time-point, providing two comparator samples for vaccine elicited boosting vs. natural decay in un-boosted subjects.

**Table 1.** Summary of COVID-19 cohort subject demographics, stratified by hospitalized (n=6) and not-hospitalized (n=18). Age, gender, ethnicity, race, recruitment population (inpatient subjects hospitalized at OHSU, Oregon Health Authority (OHA), and OHSU occupational health), and further stratifying hospitalized subjects by admitted to the intensive care unit (ICU) n=3 or not (n=3).

|  | Hospitalized |  | Total n, %<br>(n=24) | P-value |
| --- | --- | --- | --- | --- |
|  | Yes (n=6) | No (n=18) |  |  |
| Age |  |  |  |  |
| Mean (SD) | 67.0 (13.7) | 56.7 (13.2) | 59.3 (13.8) | 0.11 |
| Median [range] | 61.5 [54, 86] | 57 [28, 80] | 60 [28, 86] |  |
| Gender (n, row %) |  |  |  |  |
| Female | 1 (4) | 10 (91) | 11 (46) | 0.17 |
| Male | 5 (38) | 8 (62) | 13 (54) |  |
| Ethnicity(n, row%) |  |  |  |  |
| Hispanic | 1 (100) | 0 () | 1 (4) | 0.25 |
| Non-Hispanic | 5 (22) | 18 (78) | 23 (96) |  |
| Race (n, row %) |  |  |  |  |
| White | 5 (23) | 17 (77) | 22 (92) |  |
| Pacific Islander | - | 1 (100) | 1 (4) |  |
| Declined | 1 (100) | - | 1 (4) |  |
| Recruitment population (n, row %) |  |  |  |  |
| In-patient OHSU | 2 (100) | - | 2 (8) |  |
| OHA | 4 (17) | 10 (71) | 14 (58) |  |
| Occupational Health | - | 8 (100) | 8 (33) |  |
| Admitted to ICU (n, column %) |  |  |  |  |
| Yes | 3 (50) | NA | 3 (13) |  |
| No | 3 (50) | NA | 3 (13) |  |

### The magnitude, durability, and breadth of LLPC-derived antibodies

Forty plasma samples were collected from 24 subjects, with 15 subjects providing multiple timepoints. The VoC-RBD contained the canonical K417N, E484K, and Y501N mutations present in the currently circulating VoC. Y501N is present in the UK, SA, and P.1 strains and has been shown to increase transmission via enhanced binding to ACE-2 receptor [10]. E484K is present in 15% of strains circulating in the US (<u>CDC reporting</u> 5/21/2021). K417N and E484K are present in the B1.351 and P.1 strains [9, 25-26]. Antibody ELISA titers were below our lower-limit of detection for two asymptomatic study subjects at two separate timepoints (Fig.1 A, B) against full-length S1, RBD and VoC RBD. All but two symptomatic non-hospitalized cases had detectable antibody titers by ELISA against parental WA-1 RBD, but five symptomatic subjects had S1 ELISA titers at or below the limit of detection (at least one timepoint), and four subjects fell below the limit of detection against VoC RBD (at least one timepoint) (Figure 1A). All of the hospitalized subjects ELISA titers were above the limit of detection against S1, WA-1 RBD and VoC RBD. Overall GMT titers across antigens were highest for the hospitalized subjects (S1 858, RBD 1480, VoC 643), with symptomatic, non-hospitalized patients having lower overall GMT ELISA titers vs S1 (171), WA-1-RBD (365) and VoC-RBD (152) compared to hospitalized (Fig 1A).

**Figure 1:**
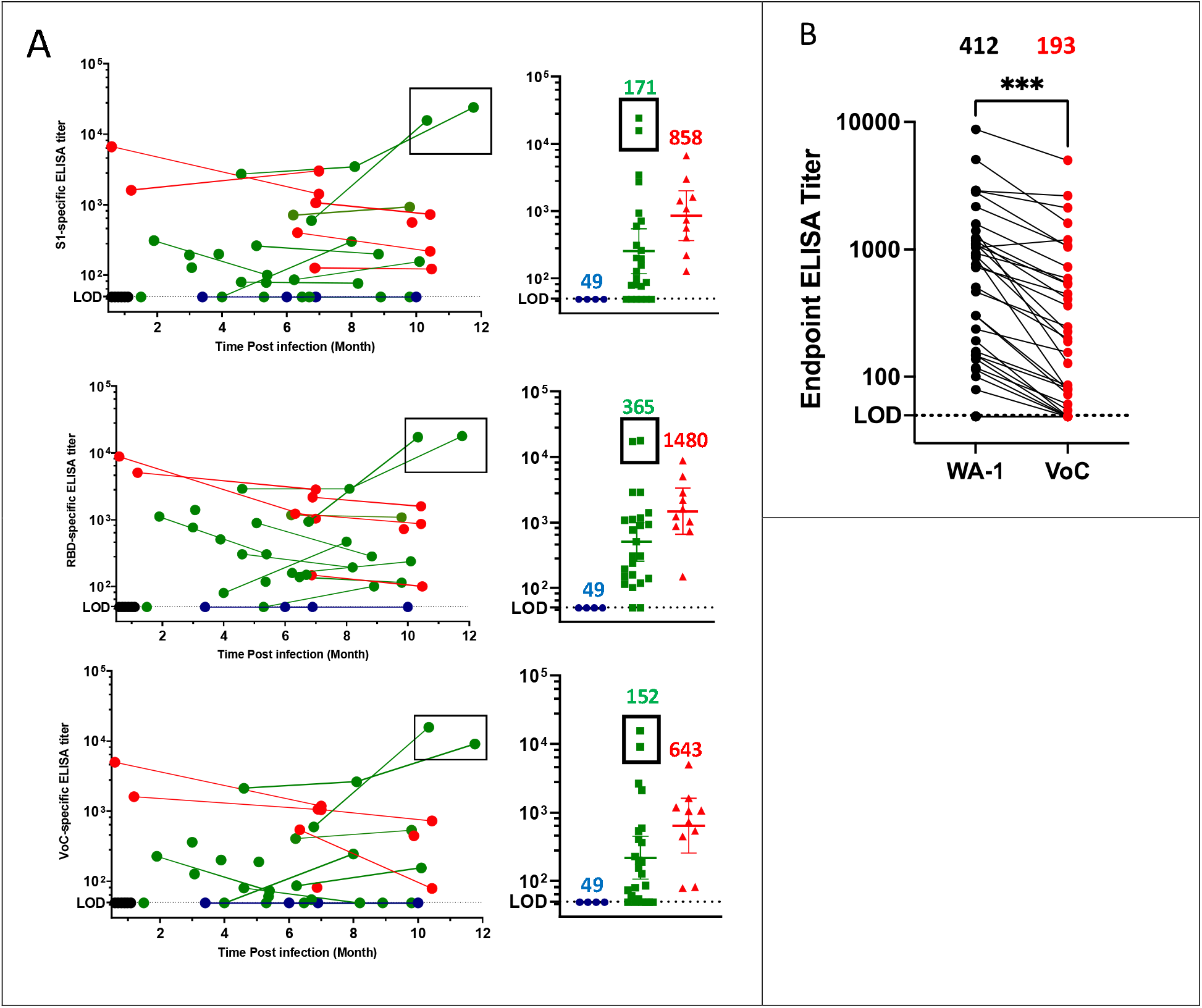
Plasma antibody titers overtime and stratified by disease severity A) Plasma ELISA titers for pre-2020 plasma (black n=6), asymptomatic (blue n=2), non-hospitalized green (n=16), and hospitalized red (n=6). Subjects who were vaccinated between sample collection are boxed out in both panels and were excluded from all statistical and data analysis. Geometric mean titers (GMT) are listed above each group. B) Comparison between ELISA titers against RBD-WA1 and VoC-RBD. Samples from subjects who received a vaccination prior to the second draw were excluded from the data analysis. Paired t-test was performed indicating significant differences between the values (p-value 0.0004) significance denoted by ^***^.

Two vaccinated subjects are included in the plotted data, with both showing a boost in ELISA titer post vaccination and both achieving the highest ELISA titers against all both RBDs in this study. The vaccinees were excluded from statistical analyses. There was a significant 2.1 -fold decrease in VoC ELISA geometric mean titers when compared to WA-1 (Fig. 1B) across all samples analyzed.

### SARS-CoV-2 specific MBC frequency remains detectable and out to 11.***75 months post-infection***

Thirty-five PBMC samples were collected from 23 subjects with 12 subjects providing multiple timepoints. In contrast to plasma Ab titers, SARS-CoV-2 S1 and RBD specific MBCs were present above the limit of detection in all subjects in this analysis, even asymptomatic subjects (Figure 2A). Moreover, SARS-CoV-2 S1 and RBD-specific MBCs remained detectable for as long as 11.75 months post-infection (Figure 2A). One subject at time-point 1 had a VoC-RBD MBC frequency below the limit of detection, however at time-point 2 this subject had detectable MBCs against RBD-VoC (Figure 2A). This is consistent with previous reports of continued germinal centers, leading to further affinity maturation over time in response to SARS CoV-2 infection [5,8,27]. When stratified by disease severity, SARS-CoV specific MBC-frequencies were closely grouped within asymptomatic and hospitalized subjects compared to non-hospitalized subjects whose frequencies that spanned the entire range from asymptomatic to hospitalized (Figure 2A). Again, two vaccinated subjects were included in the figure but excluded from analyses. Both subjects show evidence of post vaccination boost and have high overall SARS-CoV-2 specific MBC frequencies. In contrast to LLPC-derived antibodies RBD-WA-1 and RBD-VoC, MBC– derived antibodies showed little variability in their ability to recognize RBD-WA-1 vs RBD-VoC (Figure 2B) with no significant difference detected between groups.

**Figure 2:**
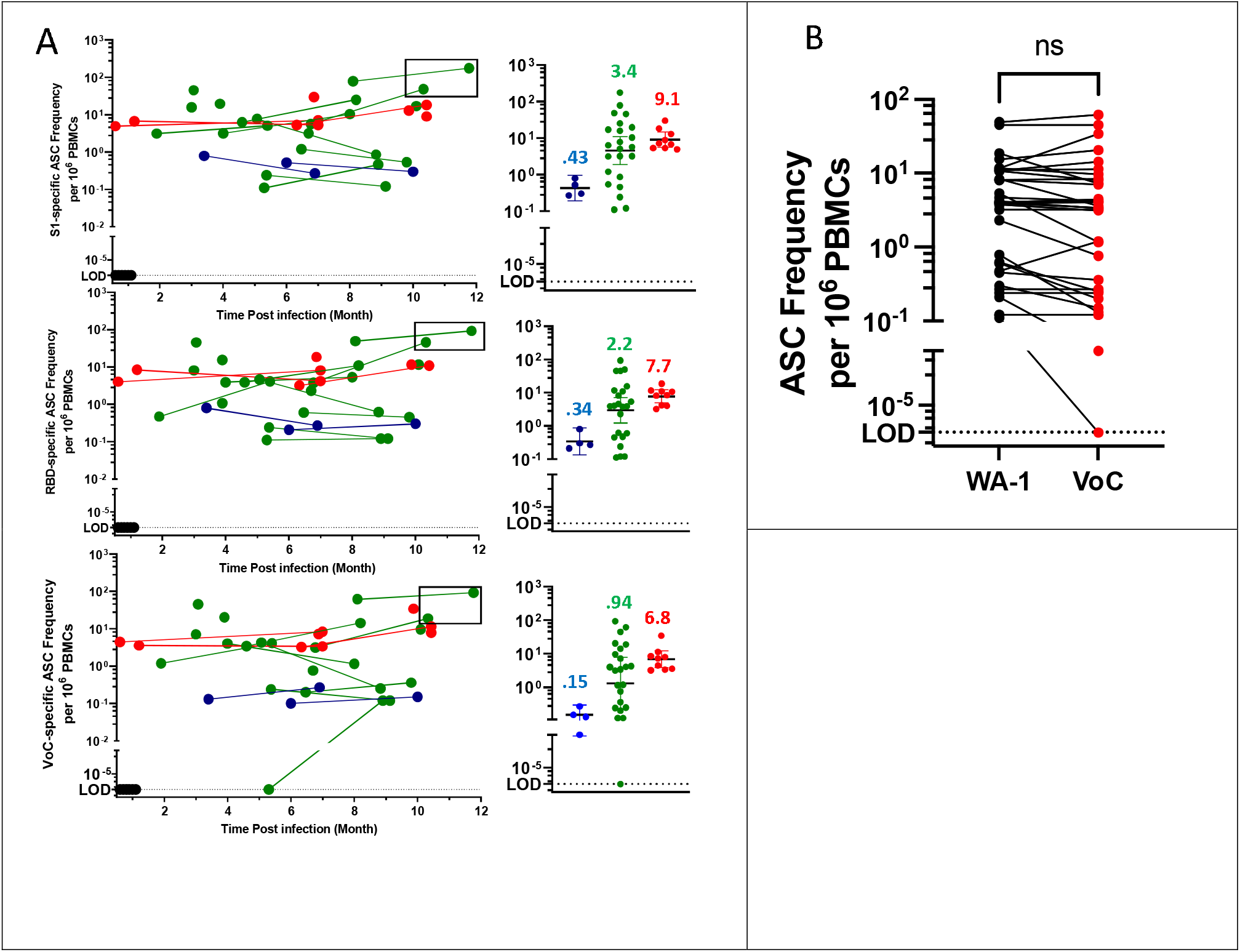
SARS-CoV-2 specific MBC frequency overtime and stratified by disease severity. A) MBC SARS CoV-2 specific frequency for pre-2020 plasma (black n=6), asymptomatic (blue n=2), non-hospitalized green (n=15), and hospitalized red (n=6). Subjects who were vaccinated between sample collection are boxed out in both panels and were excluded from all statistical and data analysis. Geometric mean titers (GMT) are listed above each group. B) Comparison between RBD-specific MBC frequency against WA1-RBD and VoC-RBD. T-test of log-transformed data, no statistically significant difference between WA-1 and VoC MBC Ab binding. Samples from subjects who had been vaccinated prior to blood draw were excluded from this analysis.

### Relationship between WA-1 and VoC ELISA titer and MBC frequency

We next examined the relationship between RBD-WA-1 specific plasma antibodies and RBD-VoC specific plasma antibodies. We observed a strong linear correlation between WA-1 and VoC RBD specific antibodies, with a Spearman correlation coefficient R^2^=0.877 p-value <0.0001, with ELISA titers against RBD-VoC approximately 3-fold lower than RBD-WA-1 across all subjects (Figure 3A), WA-1 and VoC RBD specific MBC frequencies were strongly correlated as well with a Spearman correlation coefficient R^2^=0.885 p-value <0.0001, with some samples exhibiting lower binding to VoC-RBD, but many binding equally well to both WA-1 and VoC RBDs (Figure 3B). Finally, we noted a much weaker relationship between WA-1 or VoC RBD specific ELISA titers and WA-1 (R^2^=0.346, p-value <0.0001) and VoC (R^2^=0.331, p-value 0.0004) specific MBC frequency (Figure 3C).

**Figure 3:**
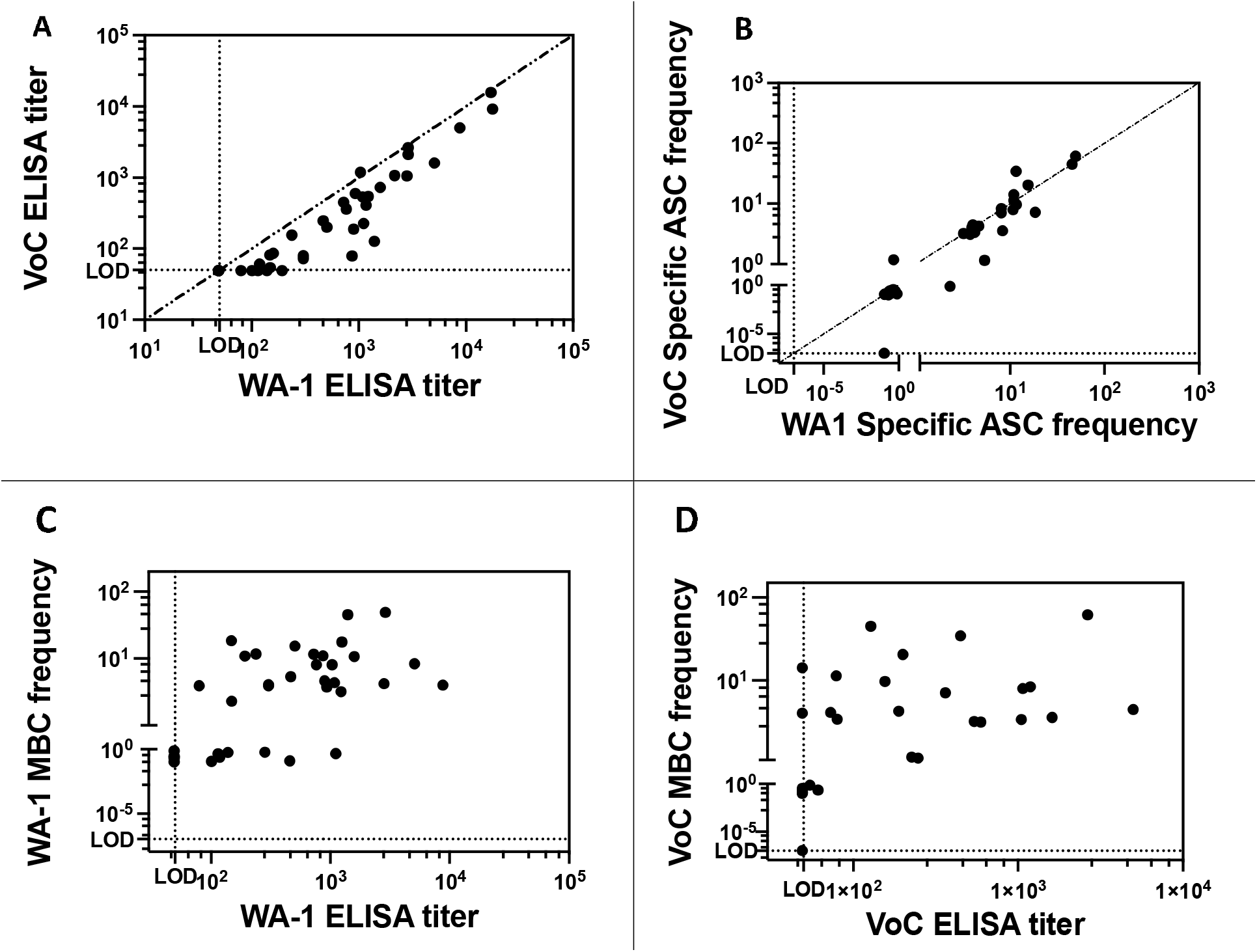
The relationship between WA-1 RBD and VoC RBD antibody binding by ELISA, WA-1 RBD and VoC RBD MBC frequency as well as the relationship between LLPC derived antibody titers and MBC frequency for each antigen. A) Correlation between RBD-VoC specific ELISA titer and RBD-WA-1 specific ELISA titer. Dotted line indicates line of identity. Spearman correlation coefficient R^2^=0.877 p-value <0.0001. B) Correlation between RBD-VoC specific MBC frequency and RBD-WA-1 specific MBC frequency. Samples from subjects who received vaccination were excluded from this analysis. Dotted line indicates line of identity. Spearman correlation coefficient R^2^=0.885 p-value <0.0001. C) Correlation between WA-1 RBD MBC frequency and ELISA titer (R^2^=.346) and between VoC-RBD MBC frequency and ELISA titer (R^2^=.331).

### The relationship between clinical/disease severity and MBC frequency

Subjects reported a variety of symptoms during acute illness (Supplemental Table 1). We found no relationship between reported symptoms and hospitalization status except for diarrhea (Fisher’s exact test, p-value <0.01). In order to test whether there was a relationship between clinical data and MBC frequencies, we conducted univariate and multivariate analyses of the relationship between age, ordinal disease severity score, and MBC frequencies. Univariate analyses found no relationship between MBC frequency, ordinal disease severity score or age (Supplemental Table 2). To complete this evaluation, we then conducted two multivariate analyses testing whether there was a relationship between age, disease score and MBC frequency. When including an interaction term between age and disease score, the model found no significant relationship between age, disease score and MBC frequency (P=0.310, Supplemental Table 3). Because the distribution of ordinal disease severity scores was largely determined by hospitalization status, we revised the model to test the relationship between age, hospitalization status, and MBC frequency, again including an interaction term for age and hospitalization status and found no significant relationship between age, hospitalization status, and MBC frequency (P=0.747).

## Discussion

Breakthrough infections and the emergence of VoC highlight the limitations of SARS-CoV-2 S specific plasma antibodies alone to recognize and protect against VoC, and work by our group and others, [9, 28-30] has shown that serum or plasma antibodies elicited by natural infection are less potent overall against several VoC. However, serologic studies do not capture the antibodies that will be secreted by MBCs when they are stimulated, expand, and differentiate into a new population of antibody secreting cells. Here we characterized the SARS-CoV-2 specific MBC derived antibodies, with three critical findings: 1) SARS-CoV-2 RBD specific MBCs can be detected in asymptomatic, seronegative, PCR-positive subjects, 2) MBC populations elicited following primary infection appear relatively stable over time, up to nearly one-year post-infection, and 3) the antibodies these MBCs are programmed to secrete recognize both parental and VoC RBDs at roughly equivalent frequencies. These results are consistent with MBC studies of other pathogens that show MBCs retain pathogen specific antibody diversity that is different from LLPC-derived Abs [11-12, 31]. Because of this additional line of antibody defense latent in SARS-CoV-2 specific MBCs, we contend there is reason for optimism regarding the capacity of vaccination, prior infection, and/or both, to limit disease severity and transmission of VoC as they inevitably emerge in the setting of ongoing transmission. The observation that the high ELISA titers were seen in previously infected subjects who went on to vaccination suggests that vaccines may be particularly effective in previously infected persons.

While SARS-CoV-2 specific antibodies were not detected in the two asymptomatic subjects on our study and 6% of our symptomatic subjects, SARS-CoV-2 specific MBCs were detectable in the peripheral blood of all subjects, including asymptomatic subjects. This result supports a recent finding that SARS-CoV-2 specific MBCs are present in the peripheral blood of subjects with no detectable SARS-CoV-2 serum Abs [11]. In this study, SARS-CoV-2 specific MBC frequency remained stable over time, sometimes even increasing, consistent with other reports [5,8,27] and consistent with evidence of extended germinal center responses that remain long after viral clearance, resulting in an increase of MBC frequency as well as an increase in diverse, high affinity MBCs. This could explain why VoC-RBD specific MBCs were detected in all but one subject, however at a later time-point VoC-RBD specific MBCs were detected in all subjects. As with plasma antibodies, we observed an increase in MBC frequency with an increase in disease severity. We hypothesize that higher antigen load correlates with higher antibody titers, similar to what has been shown in Middle Eastern respiratory syndrome (MERS),[12] as well as in SARS-CoV-2 human cohorts [13].

Study limitations include a small number of asymptomatic and hospitalized patients from whom only a subset of longitudinal samples could be collected. Moreover, our LDA approach screened antibodies specific only for S1, RBD and a specific set of known variant mutations in the RBD, while there are VoC associated mutations outside of RBD. Nevertheless, RBD has been widely identified as the most important region for neutralizing antibody epitopes, and while there are likely to be additional RBD mutations that emerge in the future, the principle that MBC-derived Ab diversity extends beyond that seen in circulating Abs is likely to hold. As more subject antibody responses are studied, more nuanced patterns than those reported here may emerge. Finally, this study evaluates the specificity of plasma IgG and IgG secreting MBCs. As SARS-CoV-2 is a respiratory pathogen, we expect IgA and MBCs programmed to secrete IgA may also play a role in protection against the variants. Additional studies that address these limitations are critically needed to further our understanding of the multifaceted roles that antibodies continue to play in controlling the COVID-19 pandemic.

## Data Availability

No additional data or external datasets

## References

1. Huang, C., et al., Clinical features of patients infected with 2019 novel coronavirus in Wuhan, China. The Lancet, 2020. 395(10223): p. 497–506.

2. Rydyznski Moderbacher, C., et al., Antigen-Specific Adaptive Immunity to SARS-CoV-2 in Acute COVID-19 and Associations with Age and Disease Severity. Cell, 2020. 183(4): p. 996-1012.e19.

3. Sette, A. and S. Crotty, Adaptive immunity to SARS-CoV-2 and COVID-19. Cell, 2021. 184(4): p. 861–880.

4. Zost, S.J., et al., Rapid isolation and profiling of a diverse panel of human monoclonal antibodies targeting the SARS-CoV-2 spike protein. Nature Medicine, 2020. 26(9): p. 1422–1427.

5. Gaebler, C., et al., Evolution of antibody immunity to SARS-CoV-2. Nature, 2021.

6. Rudberg, A.-S., et al., SARS-CoV-2 exposure, symptoms and seroprevalence in health care workers. 2020, Cold Spring Harbor Laboratory.

7. Madariaga, M.L.L., et al., Clinical predictors of donor antibody titre and correlation with recipient antibody response in a COVID□19 convalescent plasma clinical trial. Journal of Internal Medicine, 2020.

8. Dan, J.M., et al., Immunological memory to SARS-CoV-2 assessed for up to 8 months after infection. Science, 2021. 371(6529): p. eabf4063.

9. Bates, T.A., et al., Neutralization of SARS-CoV-2 variants by convalescent and vaccinated serum. 2021, Cold Spring Harbor Laboratory.

10. Slifka, M.K., et al., Humoral Immunity Due to Long-Lived Plasma Cells. Immunity, 1998. 8(3): p. 363–372.

11. Purtha, W.E., et al., Memory B cells, but not long-lived plasma cells, possess antigen specificities for viral escape mutants. Journal of Experimental Medicine, 2011. 208(13): p. 2599–2606.

12. Wong, R., et al., Affinity-Restricted Memory B Cells Dominate Recall Responses to Heterologous Flaviviruses. Immunity, 2020. 53(5): p. 1078-1094.e7.

13. Davies, N.G., et al., Estimated transmissibility and impact of SARS-CoV-2 lineage B.1.1.7 in England. 2020, Cold Spring Harbor Laboratory.

14. Tegally, H., et al., Detection of a SARS-CoV-2 variant of concern in South Africa. Nature, 2021. 592(7854): p. 438–443.

15. Wang, Z., et al., mRNA vaccine-elicited antibodies to SARS-CoV-2 and circulating variants. bioRxiv, 2021: p. 2021.01.15.426911.

16. Hajjo, R., D.A. Sabbah, and S.K. Bardaweel, Emerging SARS-CoV-2 Lineages in Middle Eastern Jordan with Increasing Mutations Near Antibody Recognition Sites. 2021, Cold Spring Harbor Laboratory.

17. Stamatatos, L., et al., Antibodies elicited by SARS-CoV-2 infection and boosted by vaccination neutralize an emerging variant and SARS-CoV-1. 2021, Cold Spring Harbor Laboratory.

18. Galloway, S.E., et al., Emergence of SARS-CoV-2 B.1.1.7 Lineage - United States, December 29, 2020-January 12, 2021. MMWR Morb Mortal Wkly Rep, 2021. 70(3): p. 95–99.

19. Cele, S., et al., Escape of SARS-CoV-2 501Y.V2 variants from neutralization by convalescent plasma. 2021, Cold Spring Harbor Laboratory.

20. Hartley, G.E., et al., Rapid generation of durable B cell memory to SARS-CoV-2 spike and nucleocapsid proteins in COVID-19 and convalescence. Science Immunology, 2020. 5(54): p. eabf8891.

21. Sokal, A., et al., Maturation and persistence of the anti-SARS-CoV-2 memory B cell response. Cell, 2021.

22. Cohen, K.W., et al., Longitudinal analysis shows durable and broad immune memory after SARS-CoV-2 infection with persisting antibody responses and memory B and T cells. 2021, Cold Spring Harbor Laboratory.

23. Pinna, D., et al., Clonal dissection of the human memory B-cell repertoire following infection and vaccination. European Journal of Immunology, 2009. 39(5): p. 1260–1270.

24. Amanna, I.J. and M.K. Slifka, Quantitation of rare memory B cell populations by two independent and complementary approaches. Journal of Immunological Methods, 2006. 317(1-2): p. 175–185.

25. Jangra, S., et al., SARS-CoV-2 spike E484K mutation reduces antibody neutralisation. The Lancet Microbe, 2021.

26. Wang, P., et al., Antibody Resistance of SARS-CoV-2 Variants B.1.351 and B.1.1.7. 2021, Cold Spring Harbor Laboratory.

27. Rodda, L.B., et al., Functional SARS-CoV-2-Specific Immune Memory Persists after Mild COVID-19. Cell, 2021. 184(1): p. 169-183.e17.

28. Chen, R.E., et al., Resistance of SARS-CoV-2 variants to neutralization by monoclonal and serum-derived polyclonal antibodies. Nature Medicine, 2021. 27(4): p. 717–726.

29. Wibmer, C.K., et al., SARS-CoV-2 501Y.V2 escapes neutralization by South African COVID-19 donor plasma. Nature Medicine, 2021. 27(4): p. 622–625.

30. Faulkner, N., et al., Reduced antibody cross-reactivity following infection with B.1.1.7 than with parental SARS-CoV-2 strains. 2021, Cold Spring Harbor Laboratory.

31. Wec, A.Z., et al., Longitudinal dynamics of the human B cell response to the yellow fever 17D vaccine. Proceedings of the National Academy of Sciences, 2020. 117(12): p. 6675–6685.

32. Winklmeier, S., et al., Persistence of functional memory B cells recognizing SARS-CoV-2 variants despite loss of specific IgG. 2021, Cold Spring Harbor Laboratory.

33. Sariol, A. and S. Perlman, Lessons for COVID-19 Immunity from Other Coronavirus Infections. Immunity, 2020. 53(2): p. 248–263.

34. Robbiani, D.F., et al., Convergent antibody responses to SARS-CoV-2 in convalescent individuals. Nature, 2020. 584(7821): p. 437–442.

35. Bilich, T., et al., T cell and antibody kinetics delineate SARS-CoV-2 peptides mediating long-term immune responses in COVID-19 convalescent individuals. Science Translational Medicine, 2021. 13(590): p. eabf7517.

36. Nelde, A., et al., SARS-CoV-2-derived peptides define heterologous and COVID-19-induced T cell recognition. Nature Immunology, 2021. 22(1): p. 74–85.

37. Oved, K., et al., Multi-center nationwide comparison of seven serology assays reveals a SARS-CoV-2 non-responding seronegative subpopulation. EClinicalMedicine, 2020. 29-30: p. 100651.

38. Marklund, E., et al., Serum-IgG responses to SARS-CoV-2 after mild and severe COVID-19 infection and analysis of IgG non-responders. PLOS ONE, 2020. 15(10): p. e0241104.

39. Masopust, D. and A.G. Soerens, Tissue-Resident T Cells and Other Resident Leukocytes. Annual Review of Immunology, 2019. 37(1): p. 521–546.

40. Stadlbauer, D., et al., SARS□CoV□2 Seroconversion in Humans: A Detailed Protocol for a Serological Assay, Antigen Production, and Test Setup. Current Protocols in Microbiology, 2020. 57(1).

41. Bates, T.A., et al., Cross-reactivity of SARS-CoV structural protein antibodies against SARS-CoV-2. 2020, Cold Spring Harbor Laboratory.

42. WHO, Novel Coronavirus COVID-19 Therapeutic Trial Synopsis, R&D Blue Print, (2020, February 18) https://www.who.int/publications/i/item/covid-19-therapeutic-trial-synopsis

