## Supplemental Table 1 for "SARS-CoV-2 specific memory B-cells from individuals with diverse disease severities recognize SARS-CoV-2 variants of concern"

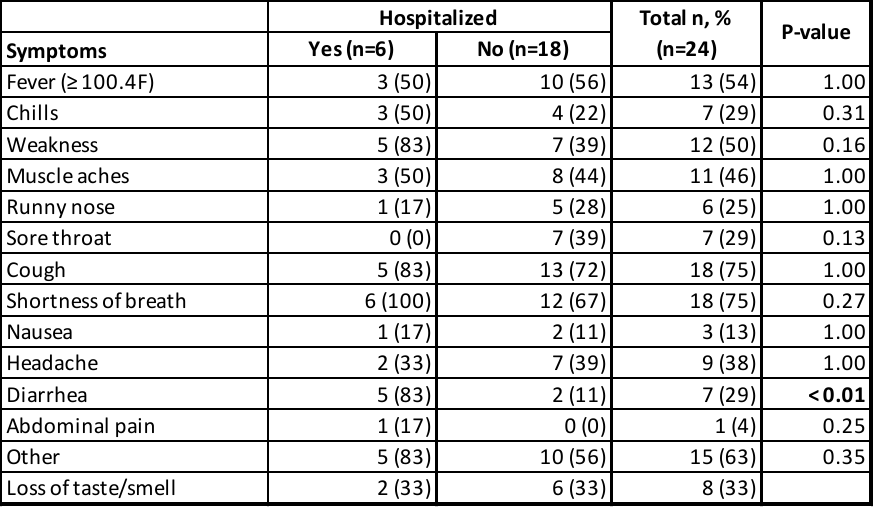
Supplemental table 1. Summary of reported symptoms experienced during acute illness Significance determined by Fisher’s exact test, p-values indicated in table.
