## Supplemental Table 2 for "SARS-CoV-2 specific memory B-cells from individuals with diverse disease severities recognize SARS-CoV-2 variants of concern"

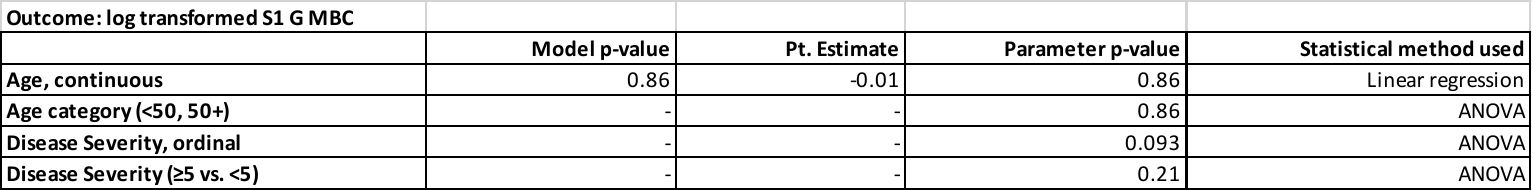
Supplemental table 2. Univariate analyses to determine relationship between MBC frequency, and disease severity score (ordinal or grouped) or age (continuous or grouped).
