## Supplemental Table 3 for "SARS-CoV-2 specific memory B-cells from individuals with diverse disease severities recognize SARS-CoV-2 variants of concern"

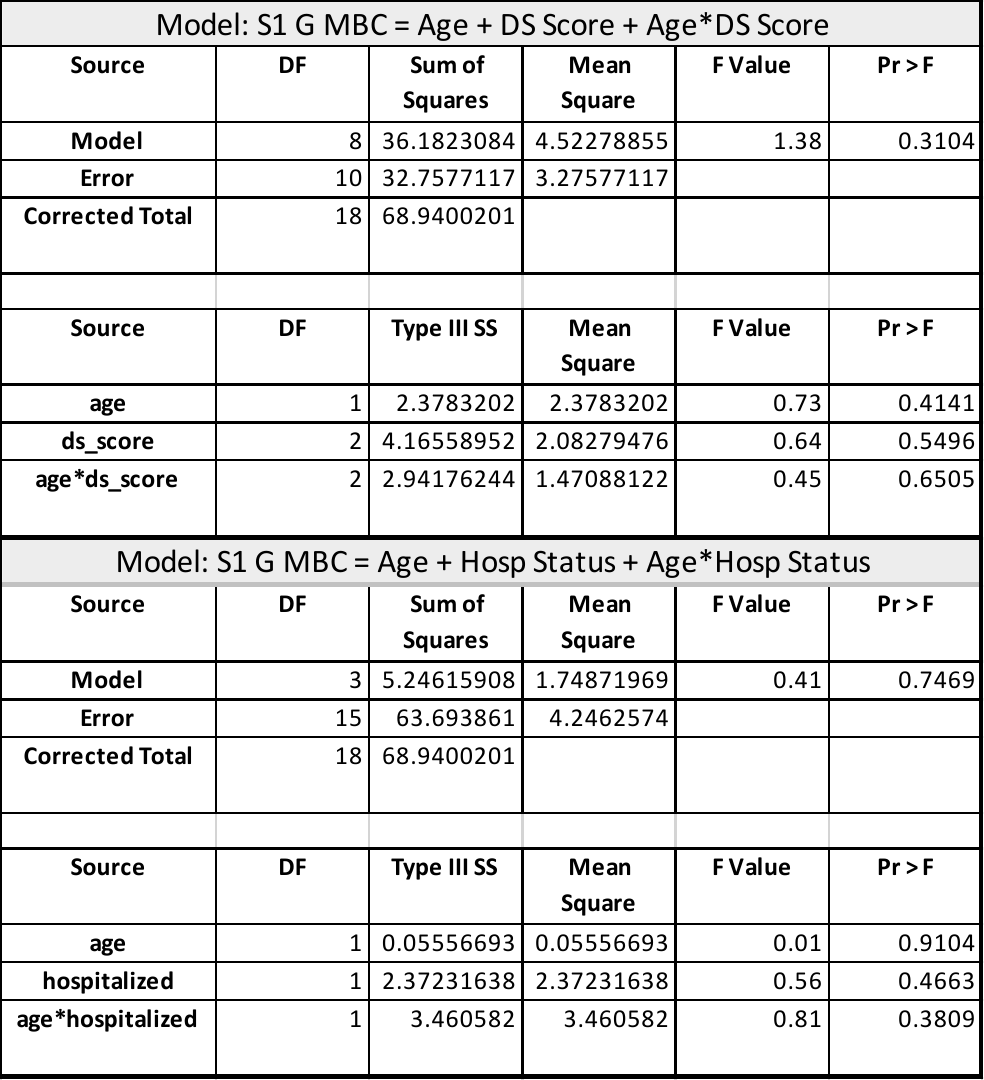

Supplemental table 3. Two multivariate analyses were run to test whether there was a relationship between age, clinical score and MBC frequency (top). Because the distribution of ordinal clinical scores was largely determined by hospitalization status, we revised the model to test the relationship between age, hospitalization status, and MBC frequency, including an interaction term for age and hospitalization status (bottom).
